# Randomized Controlled Trials of Remdesivir in Hospitalized COVID-19 Patients: A Systematic Review and Meta-Analysis

**DOI:** 10.1101/2020.08.21.20179200

**Authors:** Azza Sarfraz, Zouina Sarfraz, Marcos Sanchez-Gonzalez, Jack Michel, George Michel, Odalys Frontela, Jorge Posado, Jose Cardona, Eugonia Angueira

## Abstract

**Background:** The first cases of the coronavirus disease 2019 (COVID-19) were reported in Wuhan, China. No antiviral treatment options are currently available with proven clinical efficacy. However, preliminary findings from phase III trials suggest that remdesivir is an effective and safe treatment option for COVID-19 patients with both moderate and severe disease.

**Objective:** The aim of the present meta-analysis was to investigate whether remdesivir was effective for treating COVID-19 including reduced in-hospital adverse events, oxygen support, and mortality rates.

**Methods:** According to PRISMA reporting guidelines, a review was conducted from January 1 2020 until 25 August 2020 with MeSH terms including COVID-19, COVID, coronavirus, SARS-CoV-2, remdesivir, adenosine nucleoside triphosphate analog, Veklury using MEDLINE, Scopus, and CINAHL Plus. A modified Delphi process was utilized to include the studies and ensure that the objectives were addressed. Using dichotomous data for select values, the unadjusted odds ratios (ORs) were calculated applying Mantel Haenszel (M-H) random-effects method in Review Manager 5.4.

**Results:** Randomized controlled trials pooled in 3013 participants with 46.3% (n=1,395) in the remdesivir group and 53.7% (n=1,618) in the placebo group. The placebo group had a higher risk of mortality as compared to the intervention group with significant odds ratio (OR=0.61) (95% confidence interval of 0.45-0.82; P=0.001). There was minimal heterogeneity among the studies (I^2^=0%).

**Conclusions:** Our findings suggest that remdesivir extends clinical benefits by reducing mortality, adverse events and oxygen support in moderate to severely ill COVID-19 patients. Concerted efforts and further randomized placebo-controlled trials are warranted to examine the potency of antiviral drugs and immune-pathological host responses contributing to severity of COVID-19.

## 1. Introduction

Since the first cases of the coronavirus disease 2019 (COVID-19) were reported in Wuhan, Hubei Province, China in December 2019, the large-scale spread internationally led the World Health Organization (WHO) to declare COVID-19 as a public Health Emergency of International Concern on January 30, 2020 (1). Antiviral treatment options of proven clinical efficacy in COVID-19 infections are under investigation (2). Remdesivir is an investigational nucleotide prodrug which intracellularly metabolizes to the active nucleoside triphosphate (ATP) and interferes with viral RNA-dependent RNA polymerase activity thereby disrupting viral exoribonuclease activity (3). However, the pharmacokinetics of remdesivir within the respiratory tract of critically ill COVID-19 patients are not well known. Hospitalized COVID-19 patients with oxygen saturation ≤ 94% on room air or requiring oxygen support are eligible to receive remdesivir under the U.S. Food and Drug Administration (FDA) emergency use authorization (EUA) (4). While previous studies have reported a reduction in median time to clinical improvement, insufficient power of sample sizes limited the deductibility of clinical outcomes of remdesivir (5). Additionally, initiating remdesivir earlier in COVID-19 treatment protocols must be considered before immune-mediated epithelial damage due to elevated viral replication occurs and may reduce mortality and disease severity as observed previously in severe acute respiratory syndrome (SARS) and middle eastern respiratory syndrome (MERS) (6).

Based on preliminary reports and findings from in vitro and in vivo activity in animal models of SARS-CoV-1 and MERS-CoV, remdesivir treatment for 5 or 10 days is being administered to COVID-19 patients with comparable efficacy and safety (7–9). While most COVID-19 infections are self-limited, the largest cohort of 44,672 patients reported 14% with severe disease and 49% case-fatality rates (CFRs) among the 5% with critical disease that warrants longer hospital stays and ventilator support associated with the high burden placed on health infrastructures (10). Use of remdesivir has resulted in reduced oxygen support in a cohort with 53 hospitalized COVID-19 patients (11). Consequently, with revised recommendations suggesting uncertain efficacy of remdesivir and benefits among patients already on high-flow oxygen, mechanical ventilation or extracorporeal membrane oxygenation (ECMO), the initiation and duration of remdesivir treatment among COVID-19 hospitalized patients receiving oxygen support remains unclear (12). Given the uncertainty on the beneficial outcomes of remdesivir-treated COVID-19 patients, we aimed to examine the following differences between remdesivir and placebo groups: 1) oxygen support status at day 1 and day 14, 2) any adverse events at day 14, and 3) death from any cause at day 14.

## 2. Methods

### 2.1. Search strategy

According to PRISMA reporting guidelines, a review was conducted from January 1 2020 until 6 August 2020 with MeSH terms including “COVID-19”, “coronavirus”, “SARS-CoV-2”, “COVID”, “remdesivir”, “adenosine nucleoside triphosphate analog”, “Veklury” using Medline, Scopus, and CINAHL Plus. Quantitative primary research articles were added to the systematic review and meta-analysis. The inclusion criteria of included studies was COVID-19 infected patients aged 18 or older being treated with remdesivir or placebo. Duplicates were removed using endnote X9. We manually cross-checked the searches for authors, title, and abstract to remove duplicates.

Two investigators (AS and ZS) independently screened the titles and abstracts before reaching to a consensus to determine included studies. The third investigator (MSG) was present for any disagreements. Exclusion criteria were applied to full-texts during the final selection. A modified Delphi process was used to include studies and ensure that our objective was identified in selected studies (15). The a priori methods for conducting the Delphi process for meta-analyzing the clinical effectiveness are described in **Figure 1**. We included studies if they were randomized control trials, had an intervention arm as compared to placebo, and the endpoint of interest was clinical outcomes and mortality. Two investigators (AS and ZS) reconfirmed all data entries and checked imported data from all studies at least thrice for accuracy.

**Figure 1.**
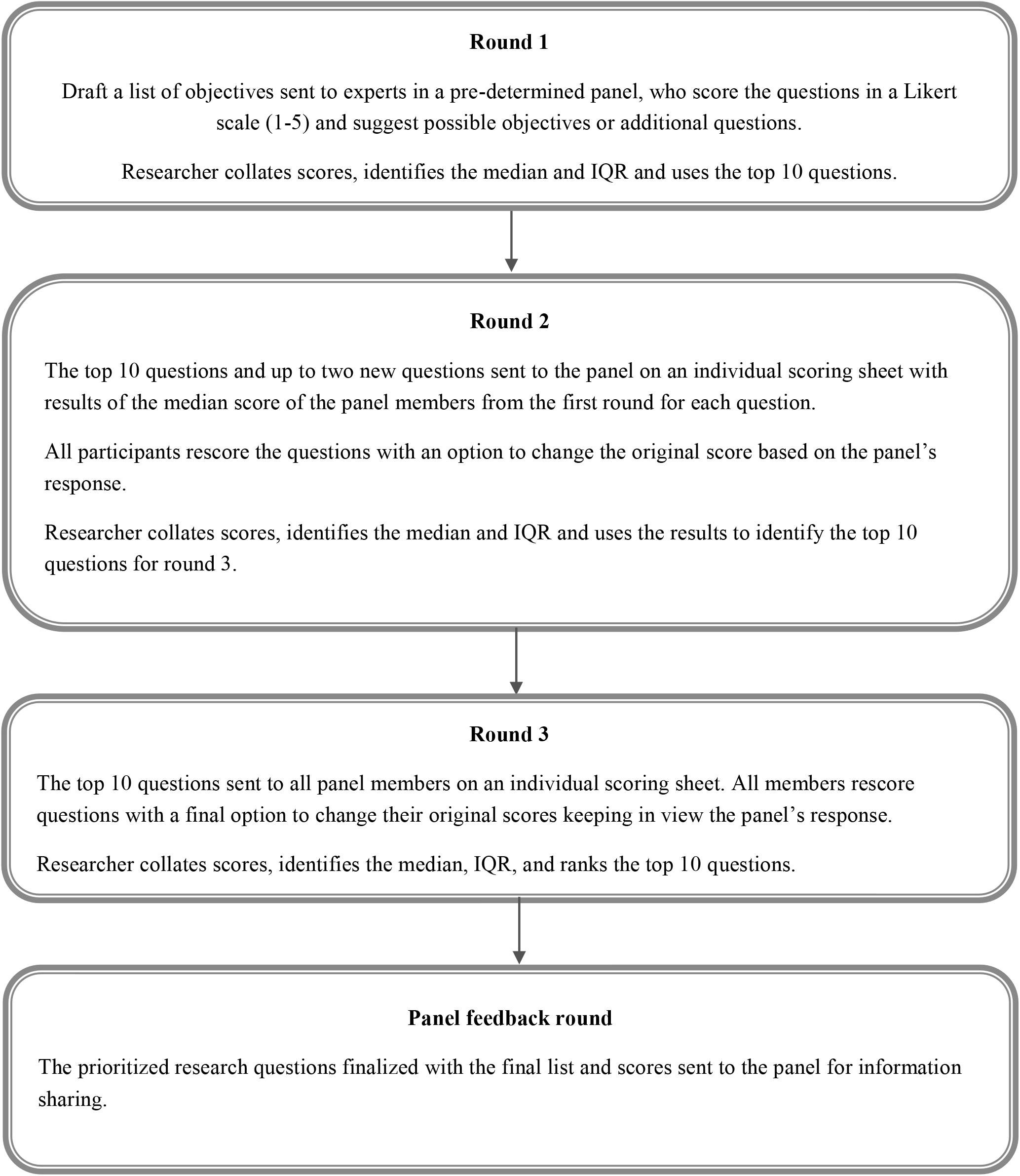
Delphi Process

### 2.2. Quality assessment

We evaluated the risk of bias for all included studies using Grading of Recommendations, Assessment, Development, and evaluation (GRADE) criteria (16). We aimed to evaluate the risk of bias associated with the selection of participants, confounding, and health outcome assessment. In doing so, we found the risk of bias of all four individual studies included for quantitative analysis using the GRADE criteria **(Table 1)**. Since less than 10 studies were included, we did not check for publication bias using funnel plots.

### 2.3. Outcomes

The primary outcomes included death from any cause at day 14. The secondary outcomes were to identify any adverse events at day 14 of the treatment and the requirement for supplemental oxygen, high-flow nasal cannula, non-invasive ventilation, invasive ventilation or ECMO at day 1 and 14. The time to recovery in days, total patients recovered, and findings of serious adverse effects among remdesivir and placebo groups were identified.

### 2.4. Data analysis

Two independent reviewers (AS and ZS) assessed the eligibility of all full-text articles; the third (MSG) arbitrated for cases to reach a consensus. The first reviewer (AS) extracted the data, and the second reviewer (ZS) validated the data extraction for all studies. The quantitative data was entered into a spreadsheet. If more than one study reported data on post-treatment outcomes, data was extracted separately for each study. We independently extracted data from the published randomized placebo-controlled trials.

By using dichotomous data for select values, summary measures namely the unadjusted odds ratios (ORs) were calculated using the Mantel Haenszel (M-H) random-effects methods. We calculated the ORs and 95% CIs for each measure evaluated in two or more studies. A meta-analysis was conducted using Review Manager V.5.4. Findings were presented using 95% CIs along with a test for heterogeneity between studies. The I^2^ index describes the inconsistency of findings across the studies in the meta-analysis reflecting the extent to which the confidence intervals of the different studies overlap.

### 2.5 Source of funding

No funding was obtained for the purpose of this study.

## 3. Results

The search process is shown in **Figure 2**. The initial screening yielded 1,242 results. After the exclusion of duplicates, 946 results were withheld for the screening of title and abstract. Consequently, 704 records were excluded due to ineligibility (reviews, editorials, non-RCTs, ongoing trials, and abstracts). Finally, after screening 242 full-text articles, only 4 studies reporting 3,013 patients (remdesivir n=1,395; placebo n=1,618) were included in the qualitative and quantitative synthesis. The major characteristics and quality assessment findings of the four included studies are presented in **Table 1**.

**Figure 2.**
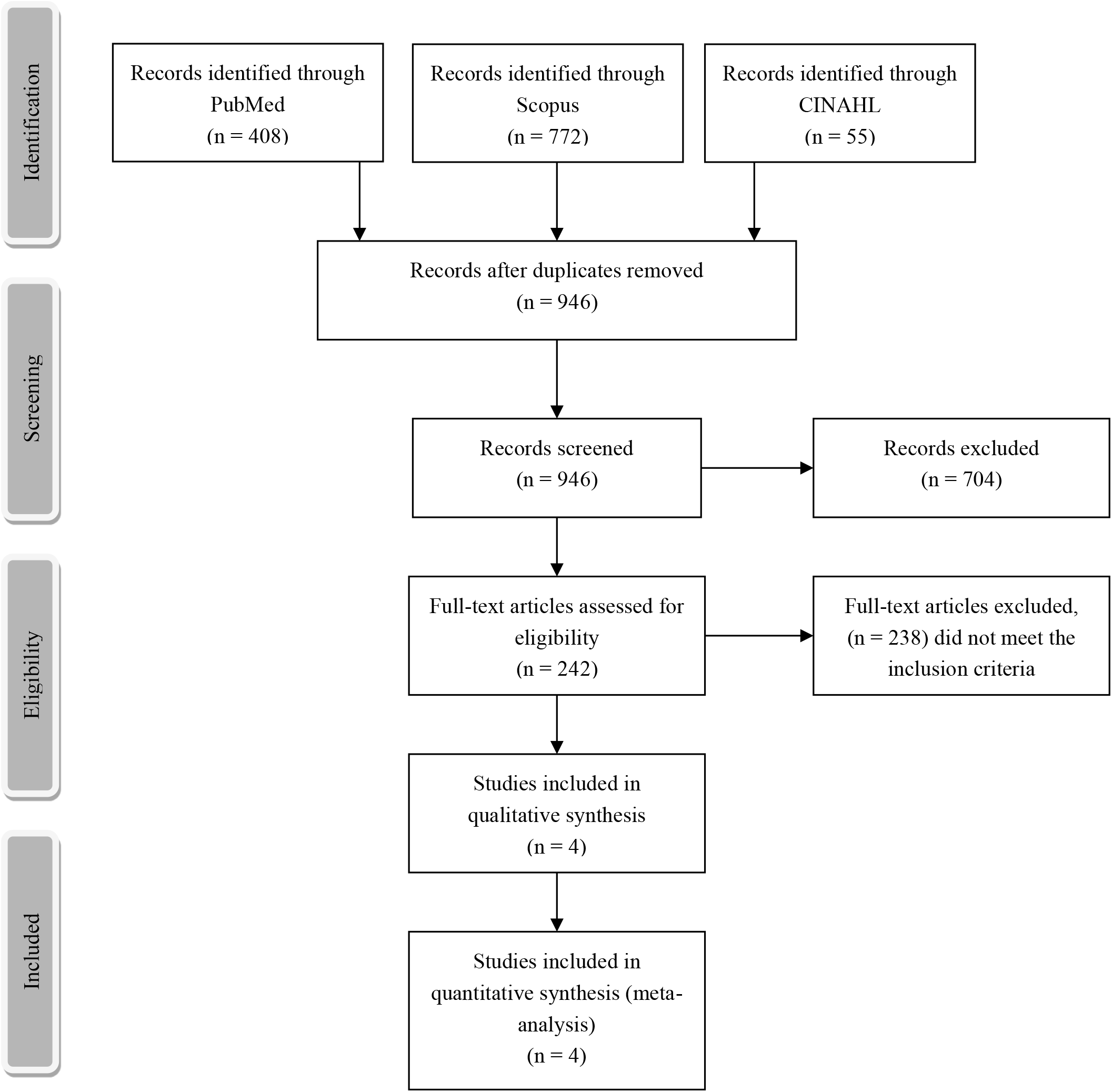
PRISMA flowchart.

**Table 1.** Study characteristics.

| Author | Groups | Age<br>(Mean<br>± SD) | Male<br>(n, %) | Hypertensio<br>n<br>(n, %) | Diabetes<br>type 2<br>(n, %) | Supplemental<br>oxygen (day<br>1)<br>(n, %) | Supplemental<br>oxygen (day<br>14)<br>(n, %) | Invasive<br>ventilation<br>or ECMO<br>(day 1) (n,<br>%) | Invasive<br>ventilatio<br>n or<br>ECMO<br>(day 14)<br>(n, %) |
| --- | --- | --- | --- | --- | --- | --- | --- | --- | --- |
| Wang<br>(18) | Remdesivir<br>(N=158) | 65.3 ±<br>12 | 89/158<br>(56%) | 72/158<br>(46%) | 40/158<br>(25%) | 129/158<br>(82%) | 61/153 (40%) | 0 (0%) | 4/153 (3%) |
|  | Control<br>(N=78) | 62.2 ±<br>12.8 | 51/78<br>(65%) | 30/78 (38%) | 16/78<br>(21%) | 65/78 (83%) | 28/78 (36%) | 1/78 (1%) | 7/78 (9%) |
| Beigel<br>(13) | Remdesivir<br>(N=541) | 58.6 ±<br>14.6 | 352/541<br>(65.1%) | 231/469<br>(49.3%) | 144/470<br>(30.6%) | 222/541<br>(41%) | 34/434 (7.8%) | 98/541<br>(18.1%) | 60/434<br>(13.8%) |
|  | Control<br>(N=522) | 59.2 ±<br>15.4 | 332/522<br>(63.6%) | 229/459<br>(49.9%) | 131/457<br>(28.7%) | 199/522<br>(38.1) | 40/410 (9.8%) | 99/522<br>(19%) | 72/410<br>(17.6%) |
| Olender*<br>(14) | Remdesivir<br>(N=312) | NA | 184/312<br>(59%) | 147/312<br>(47%) | 94/312<br>(30%) | 197/312<br>(63%) | NA | 25/312<br>(8%) | NA |
|  | Control<br>(N=818) | NA | 483/818<br>(59%) | 401/818<br>(49%) | 81/818<br>(26%) | 499/818<br>(61%) | NA | 49/818<br>(6%) | NA |
| Spinner<br>(16) | Remdesivir<br>(N=384) | NA | 232/384<br>(60.4%) | 167/384<br>(43.5%) | 156/384<br>(40.6%) | 52/384<br>(13.5%) | 9/384 (2.3%) | 0 (0%) | 1/384<br>(0.3%) |
|  | Control<br>(N=200) | NA | 125/200<br>(63%) | 81/200<br>(41%) | 76/200<br>(38%) | 36/200 (18%) | 8/200 (4%) | 0 (0%) | 5/200 (3%) |

\*Age was given in groups; remdesivir group (less than 40=11, 40 to 64 years=45, 65 years or more=44);

**Table 1. Study Characteristics (Continued).**
| Author | High-flow nasal cannula or non-invasive mechanical ventilation (day 1) (n, %) | High-flow nasal cannula or non-invasive mechanical ventilation (day 14) (n, %) | Time to recovery (days, median) | Recovered (overall) (n, %) | Serious adverse effects (overall) (n, %) | Mortality (<day 14) (n, %) | GRADE |
| --- | --- | --- | --- | --- | --- | --- | --- |
| Wang (18) | 28/158 (18%) | 13/149 (8.7%) | 21 (13-28) | 103/158 (65%) | 102/155 (66%) | 15/153 (10%) | High |
|  | 9/78 (12%) | 8/76 (10.5%) | 23 (15-28) | 45/78 (58%) | 50/78 (64%) | 7/78 (9%) |  |
| Beigel (13) | 125/541 (23.1%) | 16/538 (3.7%) | 11 (9-12) | 334/538 (62.1%) | 114/541 (21.1%) | 32/538 (6%) | High |
|  | 147/522 (28.2%) | 14/521 (3.4%) | 15 (13-19) | 273/521 (52.4%) | 141/522 (27%) | 54/521 (10.4%) |  |
| Olender (14)** | 34/312 (11%) | NA | 14 | 232/312 (74.4%) | NA | 24/312 (7.6%) | Moderate |
|  | 115/818 (14%) | NA | 14 | 483/818 (59%) | NA | 102/818 (12.5%) |  |
| Spinner (16) | 3/384 (0.8%) | 4/384 (1%) | 8 (4-13) 10 day course;<br>6 (5-10) 5 day course | 353/384 (91.9%) | 19/384 (5%) | 5/384 (1.3%) | High |
|  | 2/200 (1%) | 4/200 (2%) | 7 (4-15) | 170/200 (85%) | 18/200 (9%) | 4/200 (2%) |  |

\*\*The study compared interim data from two ongoing studies with a cutoff of fourteen days hence the time to recovery is documented until the fourteenth day.

### 3.1. Mortality at day 14 of treatment

All 4 studies reported data on mortality at day 14, and thus were eligible to be included in the meta-analysis. Compared with the remdesivir-treated group, the placebo group had higher risks of mortality (OR: 0.61; 95% CI: 0.45-0.82; P=0.001) (Figure 3a). For the sensitivity analysis, we tested if the removal of Beigel *et al*. would lead to changes in the OR and significance. After excluding this study, the results suggested that risk of mortality was still higher in the placebo group (OR: 0.66; 95% CI: 0.44-0.98; P=0.04), with homogenous findings (I^2^=0%).

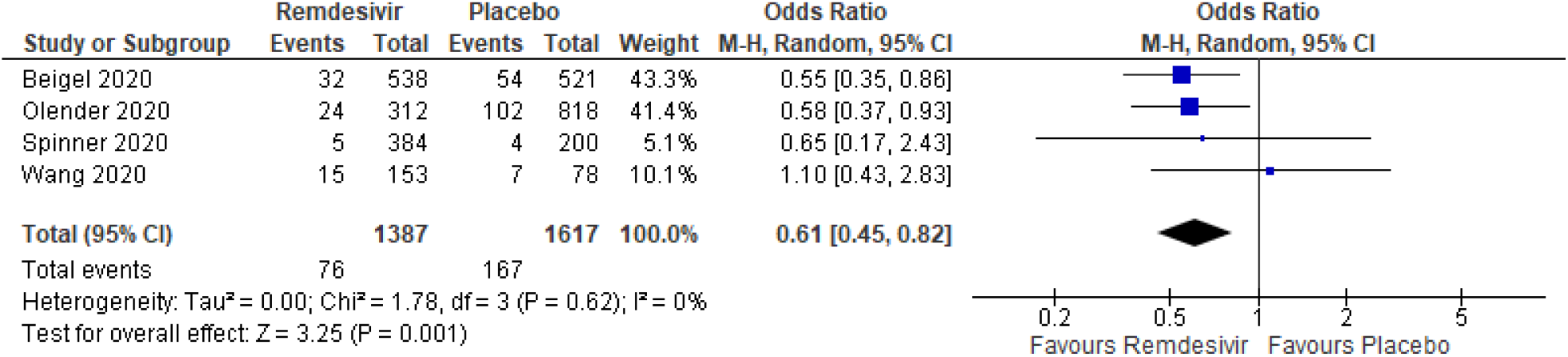
Forrest plot for mortality at day 14 of treatment.

### 3.2. Supplemental oxygen at day 1 and 14 of treatment

All 4 studies presented data of supplemental oxygen requirement at day 1 of treatment among the remdesivir and placebo groups. Using a random-effects model, we determined that the remdesivir group had similar odds as compared to the placebo group in requiring supplemental oxygen at the first day of treatment (OR: 1.03; CI: 0.87-1.23; P=0.70), with limited heterogeneity among all studies (I^2^=8%) (Figure 3b).

Three out of 4 studies evaluated the supplemental oxygen use at day 14 of treatment among the remdesivir group and the placebo group. However, there was a higher likelihood of the placebo group to require supplemental oxygen at the end of the second week of treatment (OR: 0.88; CI: 0.62-1.24; P=0.46), with no heterogeneity among the studies (I^2^=1%) (Figure 3b).

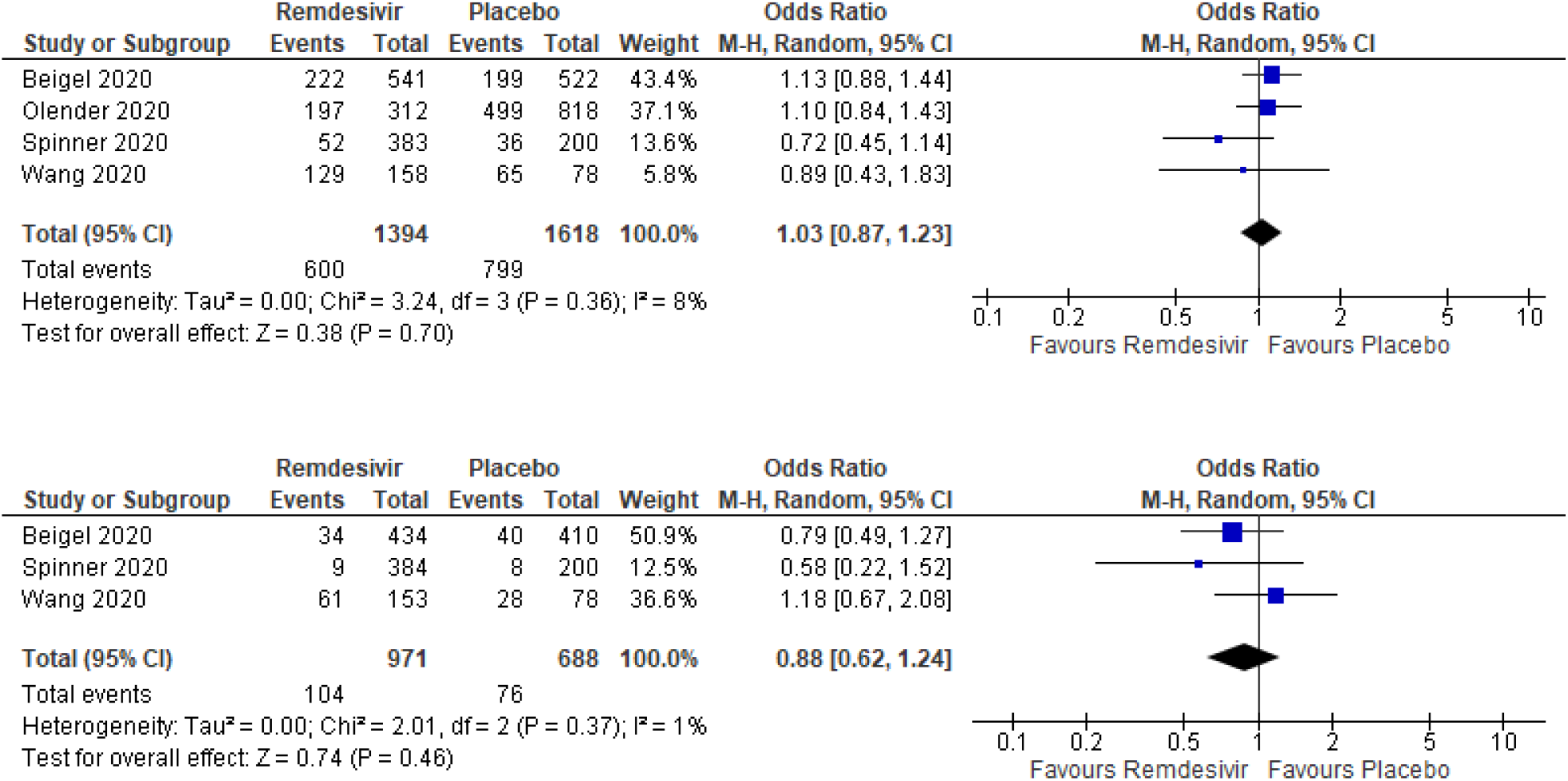
Forrest plot for supplemental oxygen at day 1 (above) and day 14 (below) of treatment.

### 3.3 High-flow nasal cannula or non-invasive mechanical ventilation at day 1 and 14 of treatment

All 4 studies presented data of high-flow nasal cannula or non-invasive mechanical ventilation required at day 1 of treatment. Patients in the placebo group as compared to the remdesivir group had high odds of requiring high-flow nasal cannula or non-invasive mechanical ventilation (OR: 0.81; CI: 0.64-1.04; P=0.10; I^2^=9%) (Figure 3c).

Three of the 4 studies presented the requirements of high-flow nasal cannula or noninvasive mechanical ventilation at day 14 of treatment. The likelihood of the placebo group was higher as compared to the remdesivir group of requiring intervention (OR: 0.90; CI: 0.53-1.53; P=0.69), with no heterogeneity among the studies (I^2^=0%) (Figure 3c).

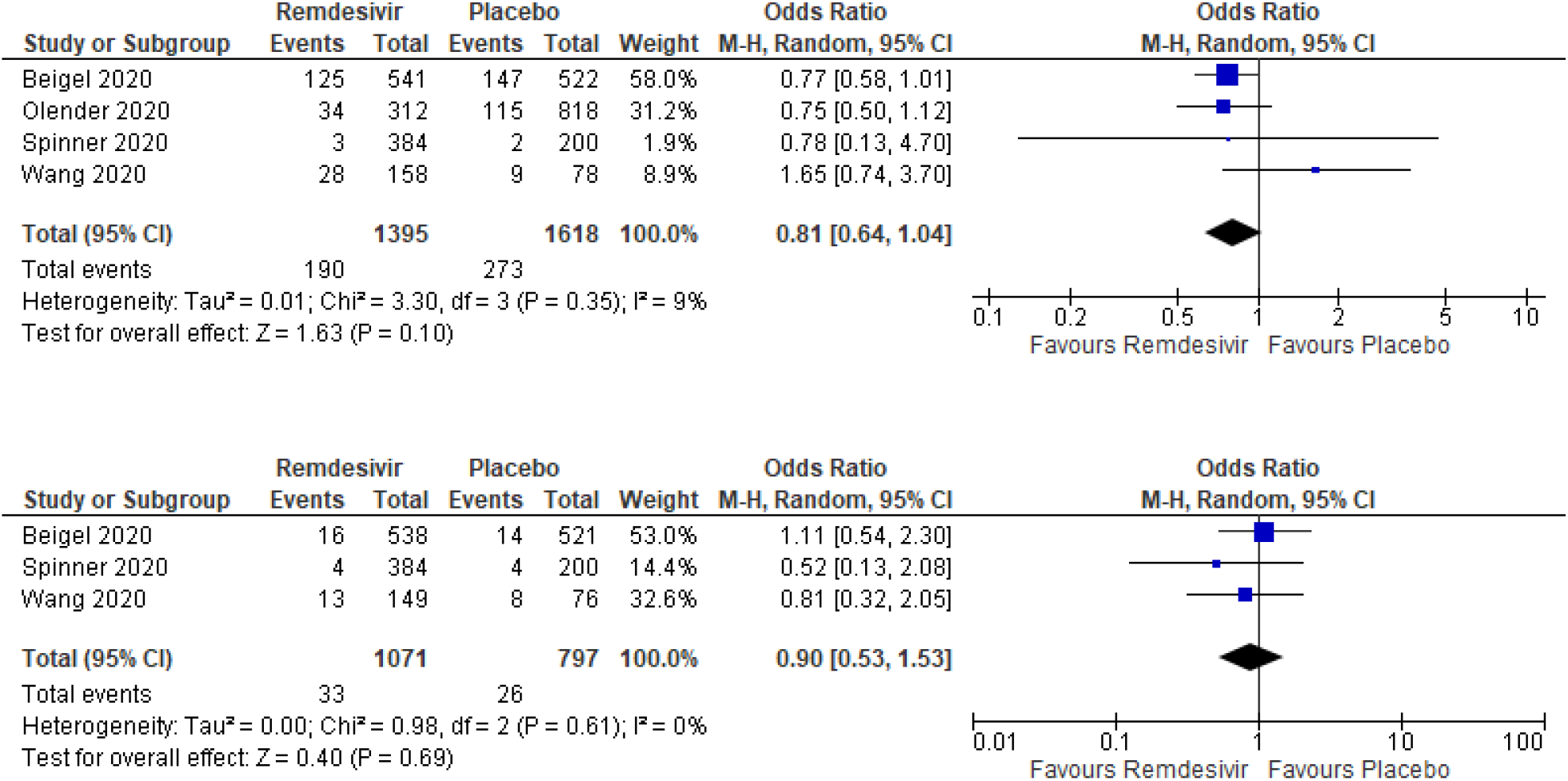
Forrest plot for supplemental oxygen at day 1 (above) and day 14 (below) of treatment.

### 3.4 Invasive ventilation or ECMO at day 1 and 14 of the treatment

Three of the 4 studies presented data of invasive ventilation or ECMO at the first day of treatment. While the difference was negligible, there was a very slight preponderance of the remdesivir group to require invasive ventilation or ECMO at day 1 of treatment (OR: 1.06; CI: 0.73-1.54; P=0.77; I^2^=28%) (Figure 3d).

Three of the 4 studies reported data on invasive ventilation or ECMO at day 14 of the treatment. Patients in the placebo group had a higher likelihood of requiring invasive ventilation or ECMO at the second week of the treatment as compared to the patients in the remdesivir group (OR: 0.39; CI: 0.13-1.14; P=0.09) (Figure 3d). There was moderately high heterogeneity among the studies included for the analysis (I^2^=62%).

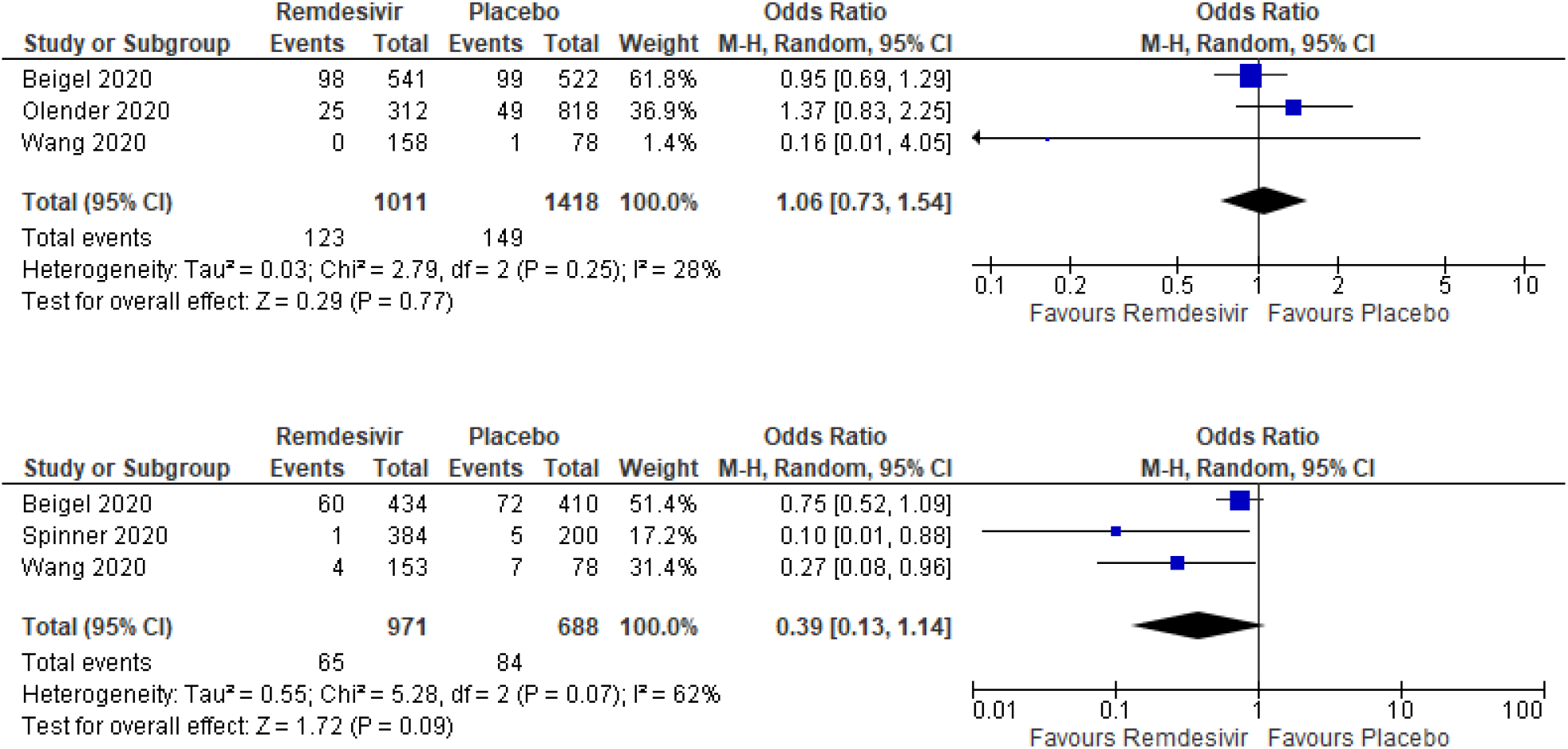
Forrest plot for invasive ventilation or ECMO at day 1 (above) and day (below) 14 of the treatment.

### 3.5 Overall Serious Adverse events after initiation of treatment

Three of the 4 studies reported data on the overall serious adverse effects initiation of treatment, and thus they were included in the meta-analyses. The placebo group had a higher risk or likelihood of presenting with adverse outcomes as compared to the remdesivir group, but with less statistical significance (OR: 0.75; CI: 0.55-1.02; P=0.07) (Figure 3e). There was mild heterogeneity between the studies (I^2^=26%).

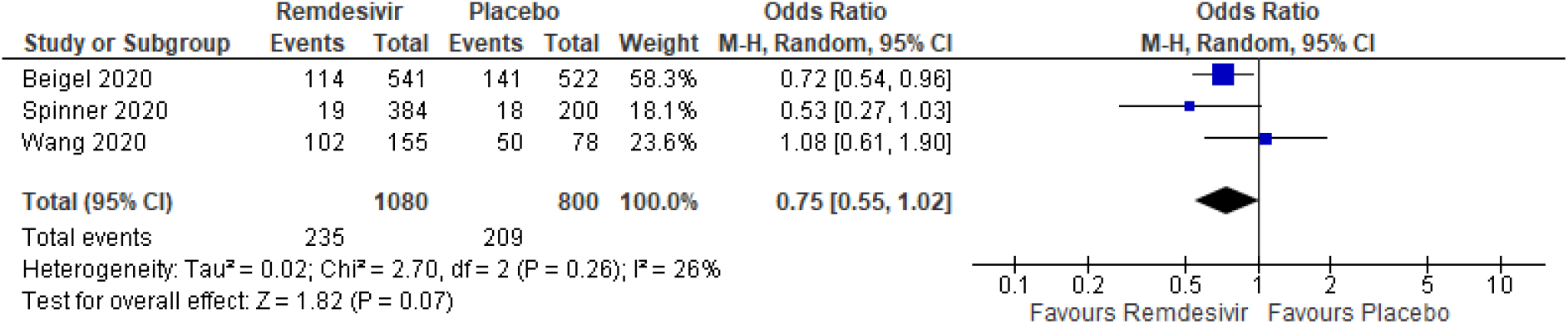
Forrest plot of serious adverse events during the entire course of the treatment.

## 4. Discussion

The purpose of the study was to comprehensively review the efficacy of remdesivir compared to placebo among hospitalized patients with moderate and severe coronavirus disease 2019. Our inclusion criteria, determined by input of all panel members, were specific for adult hospitalized COVID-19 patients treated with either remdesivir or placebo, which distinguishes the findings from other meta-analyses. Based on the analysis of four randomized placebo-controlled trials, the overall findings support the use of remdesivir to reduce oxygen support, adverse events and all-cause mortality after 5 or 10 days of remdesivir treatment (5,13,14). Overall, the mortality rate for remdesivir-treated patients with COVID-19 of the three included studies ranged from 1.3% to 10% compared to the 2% to 12.5% mortality rates of the placebo-treated patients. This findings were consistent with recent clinical data reporting positive outcomes for the compassionate use of remdesivir in moderate and severe COVID-19 patients (11,15,16).

The time to clinical recovery was significantly lower among patients who received remdesivir compared to placebo across two studies (21 days vs. 23 days and 11 days vs. 15 days). A randomized, open-label, phase 3 three-arm trial including 584 patients with moderate COVID-19 disease compared the efficacy of 5- and 10-day courses of remdesivir treatment, compared with standard care. The median time to clinical recovery across the 5 and 10-day treatment course was 6 and 8 days respectively with recovery in the standard care group being 7 days. There were observed differences in requirements of supplemental oxygen with the remdesivir group requiring less supplemental oxygen at day 14 than the placebo group with day 1 data demonstrating significant use of supplemental oxygen in the remdesivir group. While there was a very slight preponderance of the remdesivir group to require the use of high-flow nasal cannula or non-invasive mechanical ventilation at day 1, the remdesivir group had reduced likelihood of being on invasive ventilation or ECMO at day 14. Along with reduced overall oxygen support required in the remdesivir group, the all-cause mortality and any adverse events were significantly reduced in the remdesivir group in comparison to the placebo group. An analysis of 138 healthy volunteers were treated with remdesivir and it appears to have a safe clinical profile and is well-tolerated with transaminase elevation identified as the only adverse event (17). Special attention should be given to renal events, pregnancy, hypersensitivity reactions, and concomitant vasopressor use before remdesivir initiation (17).

To our best understanding, this is the first meta-analysis and systematic review of remdesivir and control groups that determines oxygen support status at day 1 and 14, any adverse events at day 14, and all-cause mortality at day 14. We synthesize various clinical outcomes of interest using statistical analyses methods that are widely applicable and relevant to key stakeholders in health care. Based on our results, implications for clinical use of remdesivir are affirmative among adult patients with COVID-19 disease demonstrated by the benefitting trends of in-hospital mortality, oxygen support status and adverse events within two weeks of treatment. Our findings synthesize results of primary and secondary outcomes of ongoing or completed clinical trials (18–20). FDA’s press release on August 28, 2020 broadened the emergency use authorization for remdesivir (e.g. Veklury) to include all hospitalized patients for the treatment of COVID-19. The scope of existing authorization is based on conclusions that remdesivir is an effective treatment option for suspected or laboratory confirmed COVID-19 patients in hospitalized settings with further trials required to explore the efficacy according to clinical stratification.

We found over 35 trials registered on clinicaltrials.gov classified as remdesivir group versus placebo group using 200 mg loading dose on the first day, followed by 100 mg intravenous once-daily doses for 5-10 days. The outcomes of the ongoing trials are to determine the time to clinical improvement, clinical status, time to hospital discharge, all-cause mortality, duration of mechanical ventilation, ECMO, supplemental oxygen, length of hospital stay, change in viral load assessed by area under viral load curve, and the frequency of adverse events.

The baseline health and disease severity were not matched in the remdesivir and placebo groups in our included studies. Additionally, the use of remdesivir in high-risk populations, e.g. elderly age, multiple comorbidity, Blacks, sociodemographic disparity, may be considered before moderate or severe COVID-19 manifestations occur (21). The most adequate time of administering anti-viral treatment is soon after the onset of disease to promote benefits, with previous reports recommending initiation within 5 or 10 days after the onset of symptoms (16,22,23). Early results based on interim data may lack generalizability, but the use of remdesivir has already obtained an approval for emergency-use authorization by the United States Food and Drug Administration (USFDA). The benefits of administrating remdesivir may outweigh the risks in hospitalized COVID-19 patients with oxygen saturations below 94%. Patients who have been intubated for a short period can also benefit from remdesivir dosage every 24-28 hours. However, limited clinical effectivity is expected among patients being mechanically ventilated.

Additionally, the next steps in finding a consensus towards remdesivir use follow the evaluation of potential short-term and long-term side effects of remdesivir taking into consideration the concomitant use of other medication. For instance, off-label use of medications such as lopinavir-ritonavir, hydroxychloroquine and immunomodulatory drugs including glucocorticoids and tocilizumab may confound reports of currently promising and beneficial outcomes of remdesivir use.

## 5. Recommendations

Reporting biases of currently published trial results may be taken into consideration. The clinical benefits ought to be predicted within all severity subgroups to confer rigorous support for clinical guidance towards remdesivir. As the world strives to overcome structural and social health care disparities, we must accentuate the underrepresentation or lack of available data interpreting the incidence, and clinical outcomes of minority groups in remdesivir COVID-19 trials (24). In a preliminary cohort study published by Grein *et al*., data of ethnicity was omitted for 53 patients (11). While the vetting for preliminary results was obtained from limited datasets the proportion of Black, Latinx, and Native Americans was around 20%, 23%, and 0.7% respectively in trials published by Beigel *et al*. and Goldman *et al*. (7,13). Additionally, while Asia’s population is roughly equivalent to 60% of the world population, Spinner et al.’s trial only consisted of 17.5% Asian participants (16). Consequently, the modest benefits in time to clinical improvement may not be generalizable to minority groups and globally due to the differences in severity, outcomes, and treatment efficacy (25). The lack of diversity is a long-standing problem that must be mandated at the administrative level by the inclusion and reporting of minority group data at government-funded research. A prioritization of populations reflecting the demographics of high-risk groups impacted by the ongoing pandemic is crucial, by expanding clinical trial sites and employing random sampling.

## 6. Limitations

Our findings were limited due to the paucity of available data between a 5-day and a 10-day course of intravenous remdesivir treatment among severe and moderate COVID-19 patients with only one randomized placebo-trial reporting these findings. All studies had open-label designs, which potentially led to biases in both patient care and reporting of data. Another limitation was the lack of corroboration of clinical efficacy with the viral loads of the patients in both groups. While the biological mechanisms of remdesivir are required to interpret the clinical efficacy, not all studies reported the viral loads in our meta-analysis.

## 7. Conclusion

Our findings provide strong evidence of clinical improvement in randomized, placebocontrolled trials of remdesivir therapy. Implications of our meta-analyses results are strong with a moderately large sample size, and randomized placebo group. Ongoing placebocontrolled trails employing larger sample sizes will remain our informative source of the outcomes and adverse events of remdesivir administered to hospitalized COVID-19 patients. Strategies must be used to enhance the potency of remdesivir while reducing the immune-pathological host responses that contribute to the infection severity. Additionally, the efficacy of 5 versus 10 days dosing of remdesivir warrants further exploration. Our findings suggest that remdesivir merits extended clinical use and may also be efficacious among non-severe hospitalized COVID-19 patients.

## Data Availability

The data that support the findings of this study are openly available in Medline or journal sites including the Lancet, New England Journal of Medicine with 10.1093/cid/ciaa1041; 10.1056/NEJMoa2007764; 10.1016/S0140-6736(20)31022-9.

https://academic.oup.com/cid/advance-article/doi/10.1093/cid/ciaa1041/5876045#:~:text=In%20this%20comparative%20analysis%2C%20by,patients%20with%20severe%20COVID%2D19.

https://www.thelancet.com/journals/lancet/article/PIIS0140-6736(20)31022-9/fulltext

